# The role of telemedicine towards improved sustainability in health care and societal productivity in Turkey

**DOI:** 10.1101/2024.03.20.24304591

**Authors:** Figen Özen, Alptug H. Kaynar, A. Kubilay Korkut, Melike Elif Teker Açıkel, Z. Dilsun Kaynar, A. Murat Kaynar

## Abstract

The COVID-19 pandemic increased utilization of telemedicine for diagnosis and treatment. While telemedicine is not the panacea for the increasing health care burden, it can alleviate the problem. Here, the hypothetical impact of delivering telehealth care to patients in a busy tertiary cardio-vascular clinic in Istanbul, Turkey, is examined. Additionally, the potential environmental and societal ramifications of telemedicine are also examined.

Demographics, health care costs, wages, productivity, and patient-specific data were exploited to develop a hypothetical telemedicine framework for the Turkish health care landscape. Specifically, the distance traveled and travel time to receive care using real-life location of the clinics and patients addresses seeking care are tabulated.

Data from August 3, 2015 to January 25, 2023 resulted in 45,602 unique encounters with 448 unique diagnoses recorded for the patient encounters. The patients in the top 5% of the most common diagnoses traveled 23.82 ± 96.3 km to reach the clinics. Based on our model and the related literature that telehealth care for chronic diseases is not inferior to face-to-face care, 656,258 km would have been saved if all patients were to take the first visit in person followed by telemedicine visits. The travel-associated carbon footprint and wage losses for in-person care in lieu of telehealth appointments is calculated and it was observed that exploiting telemedicine could have saved approximately 30% carbon footprint and prevented approximately $878,000 wage loss. As a result, it is found that application of telemedicine could ease the burden on patients, environment, increase access, and prevent the wage losses caused by unnecessary hospital visits.

## 1. Introduction

The COVID-19 pandemic forced a change in the way we practice medicine, leading to an increased use of telemedicine in diagnosing and treating diseases. In this study, we examined the hypothetical savings in travel distance and time for hospital visits in Istanbul, Turkey enabled by healthcare using telemedicine in the form of synchronous video conferences. Our study used busy tertiary cardiology and cardio-vascular surgery clinics in Istanbul as a model to argue for the potential positive environmental, economic, and societal ramifications of telemedicine in settings that are similar to our model where specialized care is concentrated in certain clinics in a populous city only accessible via often traffic-laden travel routes.

We used datasets for demographics, health care costs, wages, productivity, and data of patients of the above-mentioned clinics to develop a hypothetical framework for the Turkish health care landscape as we build a case for the use of telemedicine. Specifically, we obtained the distance and time expended to receive care at the clinics by using the addresses of the clinics and patients seeking care. The calculated distances and travel times allowed us to calculate the carbon footprint and wage losses associated with traveling to seek in-person care in lieu of telemedicine appointments. Recent literature has demonstrated the non-inferiority of outcomes in telemedicine treatment compared with face-to-face treatment for patients with peripheral arterial and venous diseases [1]–[3].

Our findings, based on the model we developed using the existing patient-level data suggests (1) the plausibility of remote care and follow-up of patients suffering from cardio-vascular diseases, (2) associated benefits to the environments, and (3) savings in wages.

### 1.1. Background

The right to health care was recognized as a human right at the 1966 International Covenant on Economic, Social and Cultural Rights. By this principle, governments have an obligation to deliver accessible healthcare to their constituents. However, there is a cost to deliver and receive health care. With ever growing health care costs and its impact on the gross domestic product (GDP) of each country, governments are struggling to find ways to fulfill this promise (Fig. 1) [4].

**Figure 1:**
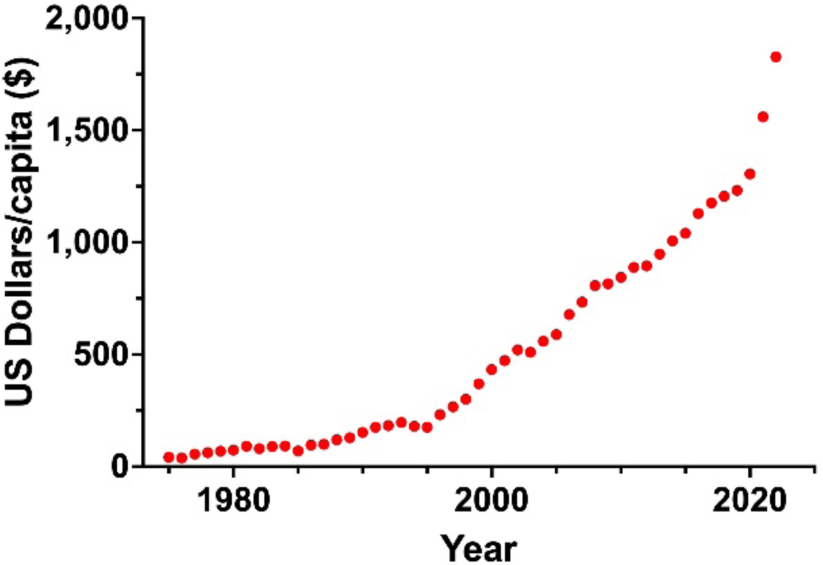
Cost of health care over time in Turkey.

Rising health care cost is especially a growing concern in low and middle-income countries (LMIC), such as Turkey (Fig. 2)^2^ [5], [6].

**Figure 2:**
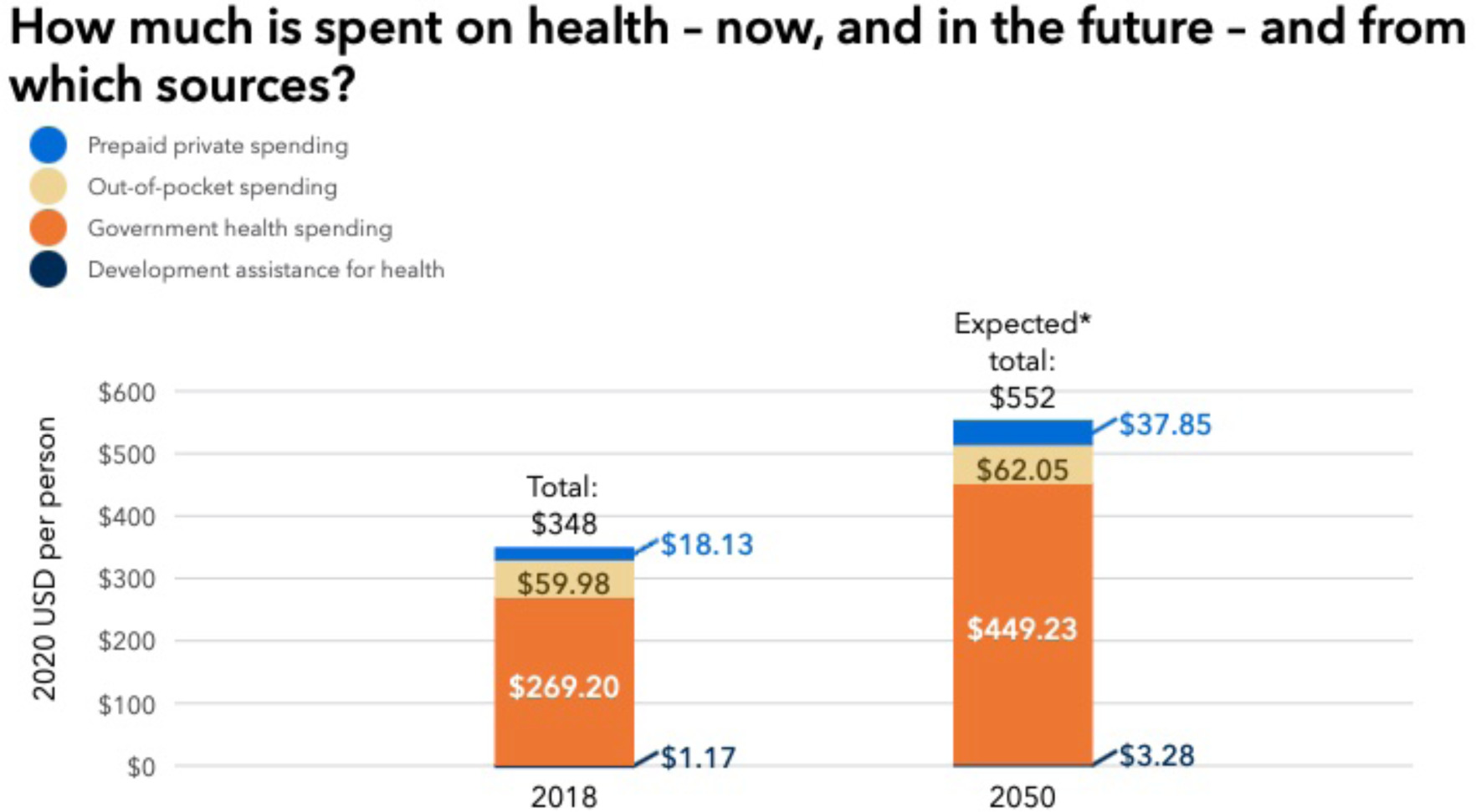
The predicted cost of health care in Turkey from 2019 to 2050, rising from a total of $378 in 2019 to $694 in 2050.

The steady growth in the Turkish population puts ever-increasing strain on its limited budget health. In a recent work from Turkey, it was reported that the hospitals run by the Ministry of Health in Turkey allocate doctor appointments for every 7.5 min underlying the strain on the system, providers, and patients [7]. As such, Turkey’s population density will guide policy makers toward more innovative care models (Fig. 3) [8].

**Figure 3:**
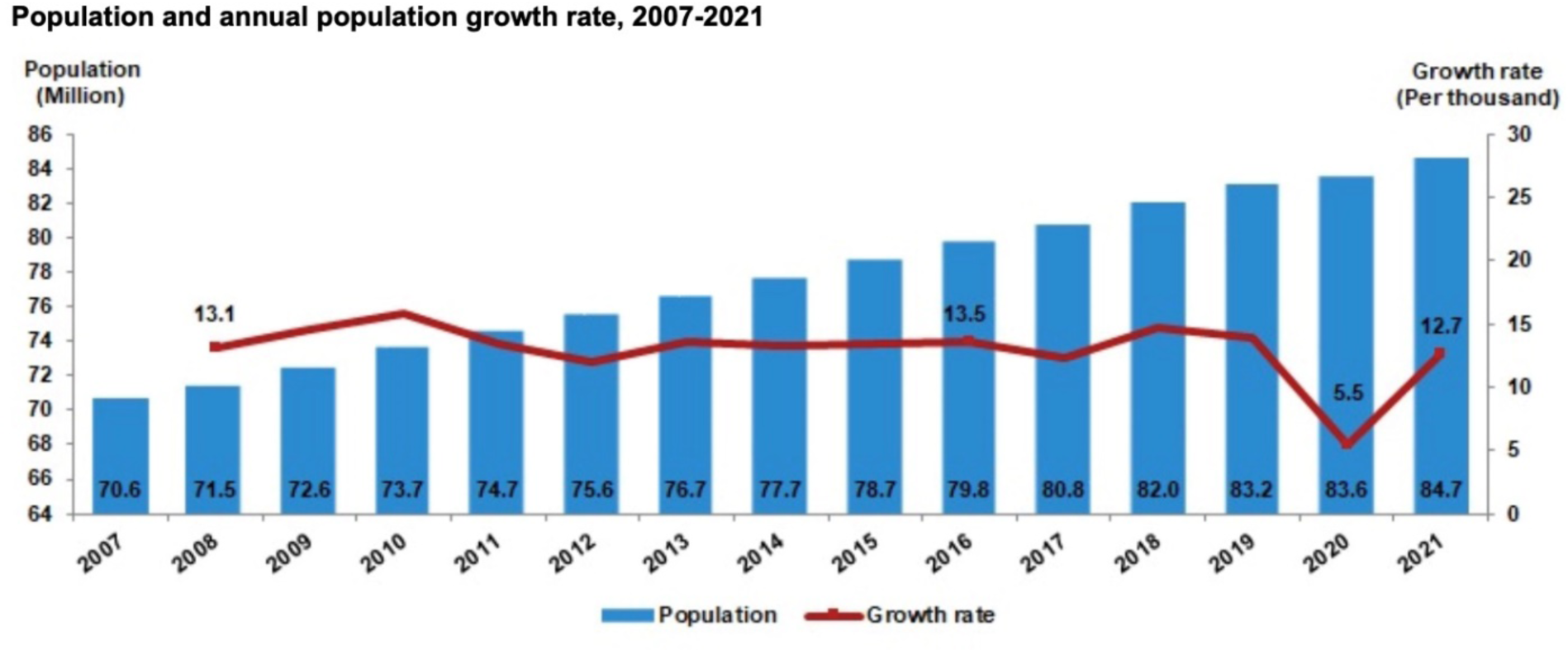
Annual population growth rate in Turkey increased to 12.7 per thousand in 2021 from 5.5 per thousand in 2020.

To compound the difficulties posed by growing health care costs, the exodus of the health care workforce into other careers following the COVID-19 pandemic has put additional strain on health care systems globally [9]–[14].

While the pandemic was disruptive, it also accelerated the integration and acceptance of innovative technologies in delivering care [15], [16]. One such technology is telemedicine, which allows health care providers to care for patients without an in-person office visit. As the concept of telehealth is taking root, clinical trials using telemedicine and integration with artificial intelligence are also increasing [17]. In addition, the incorporation of remote patient monitoring technologies will accelerate the wide use of telemedicine [18].

There are published and ongoing clinical trials for personalized telemedicine around cardiovascular health [19]. A recent work compared the use of telemedicine to in-person doctor visits among patients with congestive heart failure (CHF). Among 850 patients with CHF, a hybrid telemedicine care model was not inferior to in-person visits for mortality [20]. In a trial among 1,119 patients with CHF, telemedicine-based management emergency service for high-risk HF patients proved to be safe and reduced unplanned hospitalizations [21]. Another trial showed decreased emergency medicine department admissions among patients with CHF if they were followed up by a tele-pharmacist [22]. Based on the clinical trials, there is an important and effective role for telemedicine in health care [23]–[25].

In the case of Turkey, telemedicine acceptance was fast during the Covid pandemic, yet for any new system to be successful it requires the capacity for expansion, financing, policies, governance, and partnership [26], [27]. The hypothetical benefits of adopting telemedicine presented in this work could inform and motivate future policies in this space.

## 2. Materials and methods

### Declaration

The study was conducted in accordance with the Declaration of Helsinki. The study was approved by the Institutional Review Board of the Haliç University, Turkey (September 26, 2023; No. 212).

In this work, we calculated the average distance and time spent reaching to health care facilities in Istanbul, Turkey as the primary outcomes and associated carbon footprint and wage losses as secondary outcomes.

### 2.1. Travel distance and time

Travel distance and time were obtained by web scraping Google Maps travel time and distance data. The patient address dataset and patient demographical data were obtained from three clinics (Haseki Merkez, Haseki Sultangazi, and Haseki 29 Mayıs) in Istanbul serving a large catchment area (Fig. 4). Using Selenium browser automation, each patient’s address was matched to their respective branch of the Haseki hospital system in Istanbul, Turkey. This matched patient address-hospital pair was inserted into Google Maps and the resultant travel time and distance data, obtained on the “driving” setting, was recorded. The timing would have been difficult to predict using the public transportation option as the times using public option are not predictable. The timing of the Google Map data extraction was performed after hours (5:00 PM until 9:00 AM EST, 9 AM until 5 PM UTC + 03:00) to minimize the impact of an inaccurate representation of traffic on the time calculations.

**Figure 4:**
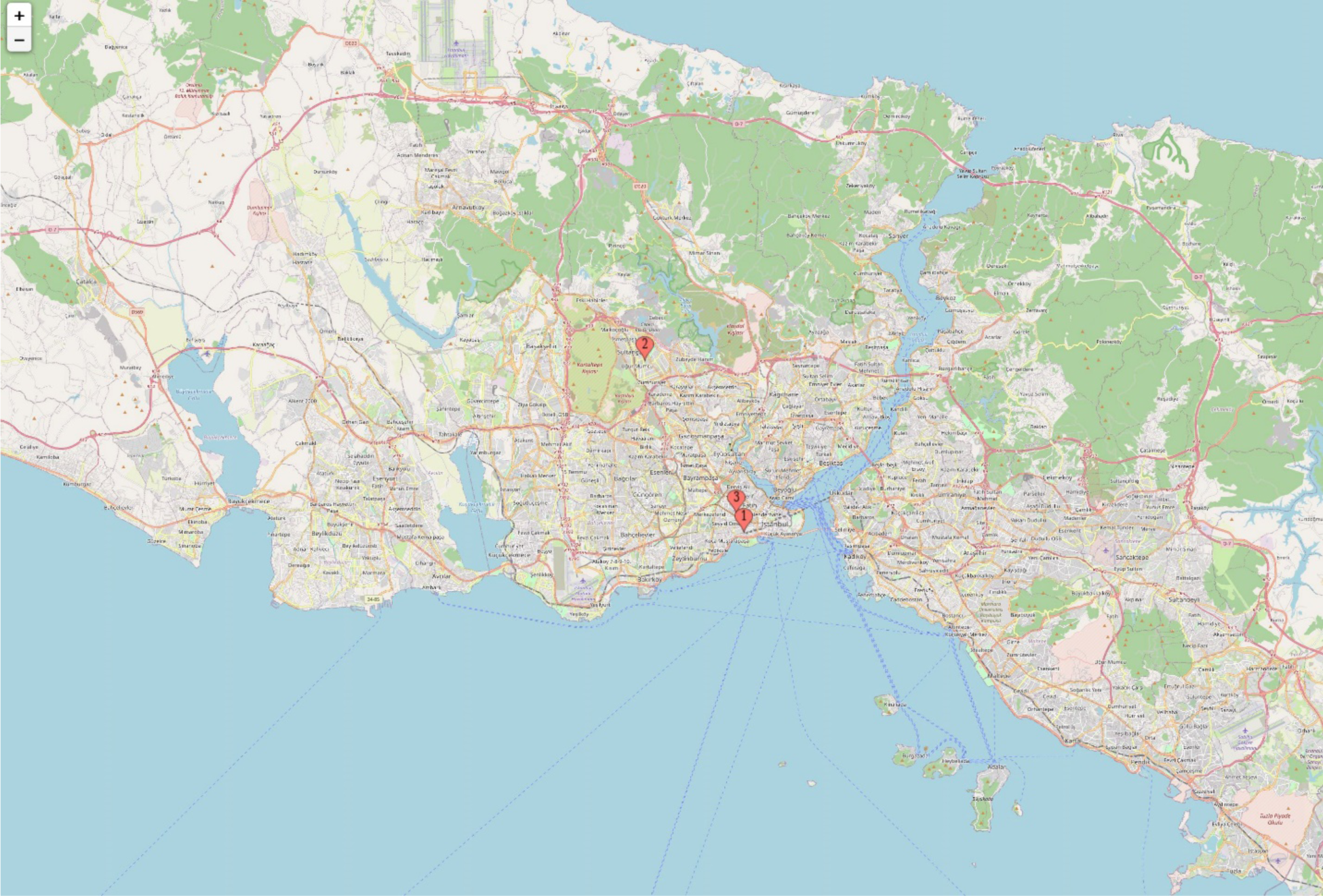
The patient address dataset and patient demographical data were obtained from three clinics (Haseki Merkez, Haseki Sultangazi, and Haseki 29 Mayıs) in Istanbul serving a large catchment area.

### 2.2. Carbon footprint of traveling to health care facilities and telemedicine alternatives

The carbon emissions were calculated based on assumed modes of transportation with models varying from personal vehicles to public transportation as well as the carbon emission from synchronous videoconference-based telemedicine. The carbon emission data per km is derived from the Global Change Data Lab [28] and the telemedicine carbon footprint data is obtained from the work by Obringer et al. [29]. In the example of in-person visit, a 10-minute doctor-patient interaction is assumed. Similarly, for telemedicine, the digital footprints are calculated for a 10-minute doctor-patient telemedicine conferencing and it is assumed that on the average one large image is exchanged for diagnosis.

### 2.3. Economic impact of traveling to health care facilities

The lost wage cost is derived from the nation-specific wage and employment data from the International Labor Organization (ILO) for the year 2021 and the associated labor wage data for that year. The retirement age in Turkey is 58 for women and 60 for men. The two-way travel time and the additional estimated 10 min for physician interaction is multiplied by the hourly wage.

## 3. Results

### 3.1. The distance and time to travel to health care facilities

The dataset included 64,999 unique patient visits from August 3, 2015 to January 25, 2020 to the three health care facilities in Istanbul (Haseki Merkez, Haseki Sultangazi, Haseki 29 Mayıs). After correcting for missing or inaccurate address information, we had a complete address dataset of 45,602 unique patient encounters.

There were 448 unique diagnoses recorded for the patient encounters and we decided to examine the top 5% of diagnoses (Fig. 5).

**Figure 5:**
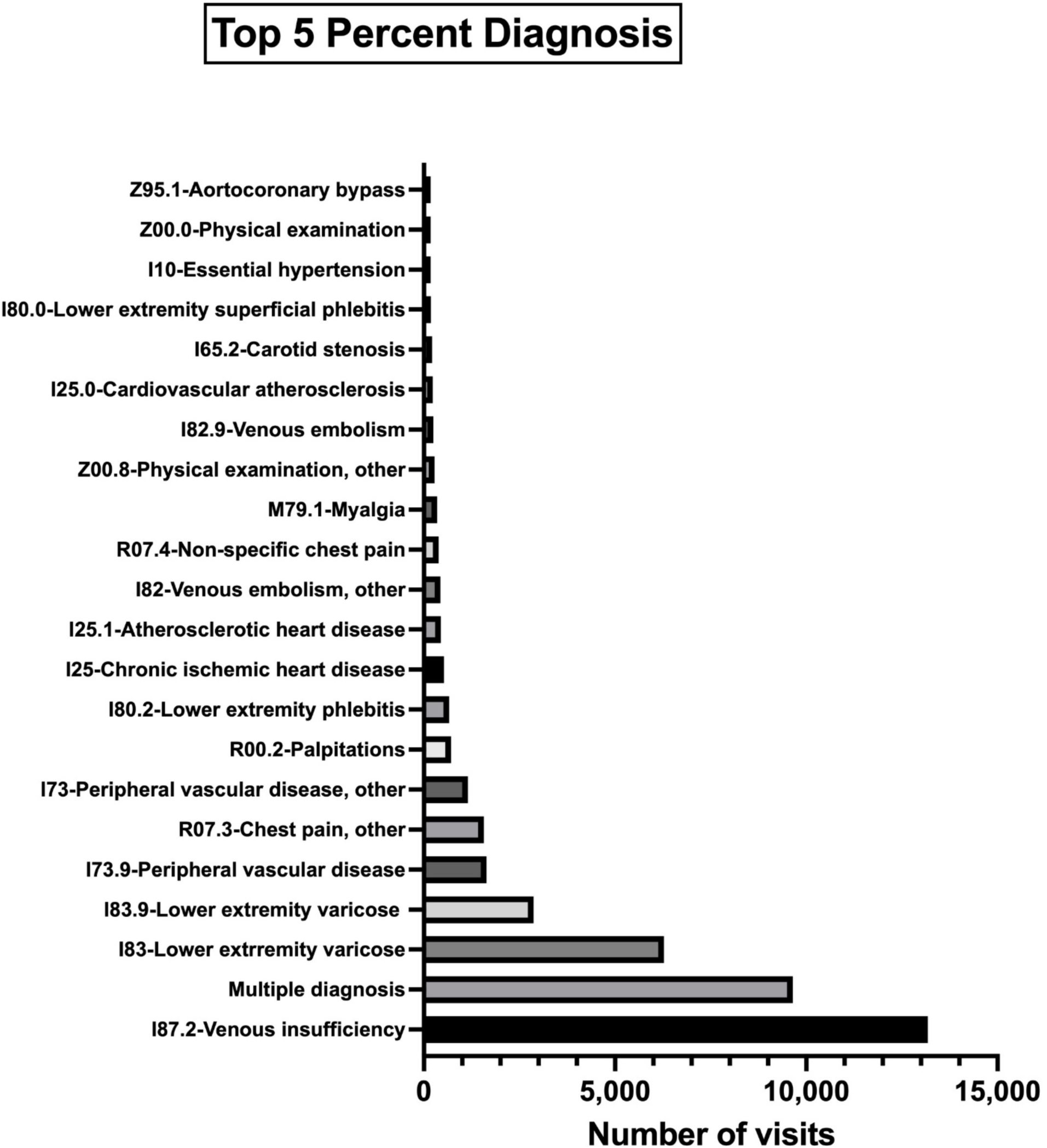
The top 5% of diagnosis for the visits, mainly around cardiovascular diseases.

Patients traveled on average 23.82 ± 96.3 km to reach the health care facilities (Fig. 6 and 7). As shown in Fig 7, there would be a total of 656,258 km saved if all patients were to take the first visit in person followed by telemedicine for subsequent visits making it a potential saving. Our analysis assumes that the visits that follow the first in-person visit could be replaced with synchronous video conferencing. This is because our data showed that, for the diagnoses included in our analysis, 98.9% of the visits subsequent to the first were follow-up visits that did not require any in-person imaging, wound care, or surgical intervention that would be impossible to do virtually.

**Figure 6:**
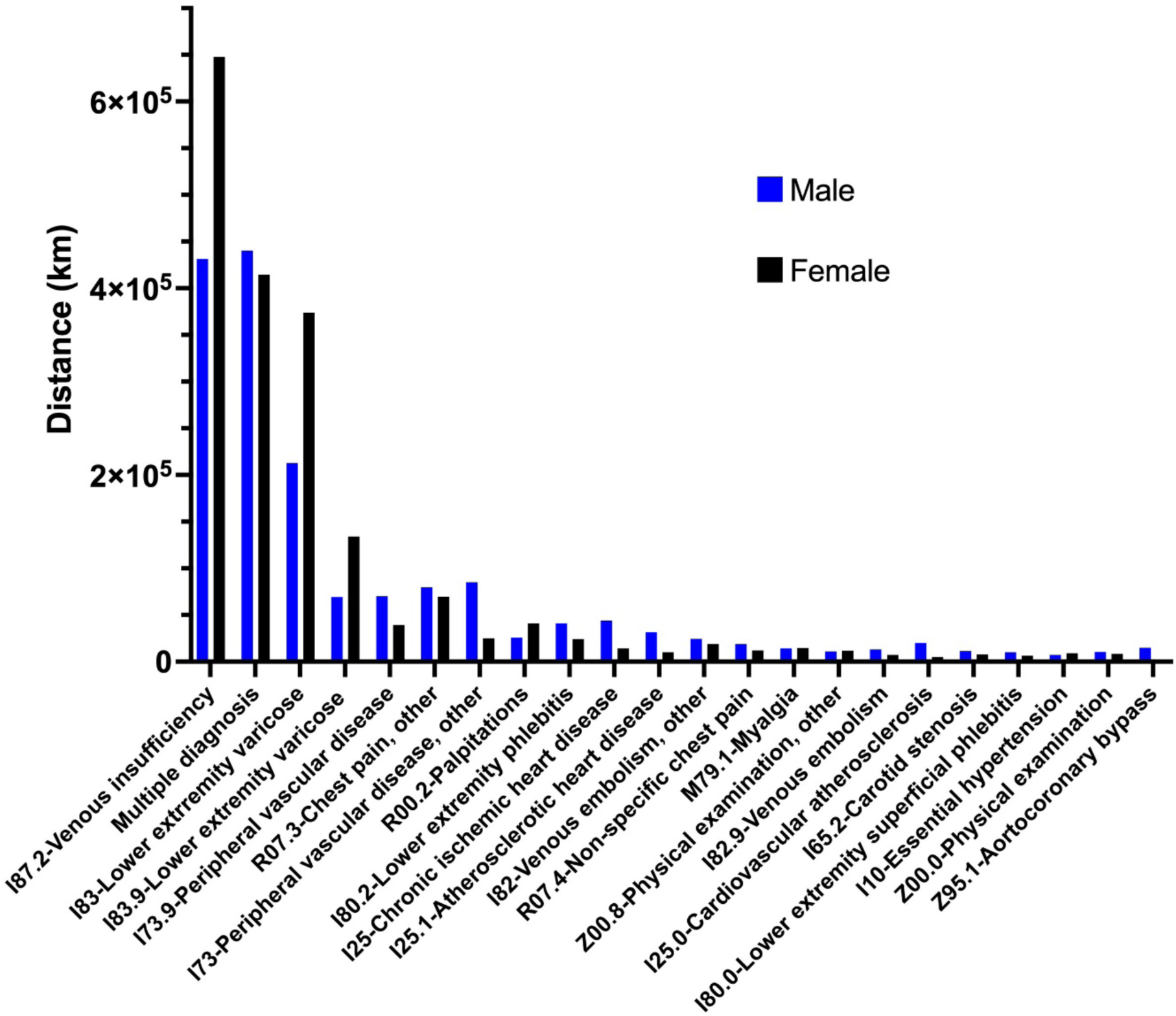
Distance traveled by the top 5% diagnosis groups.

**Figure 7:**
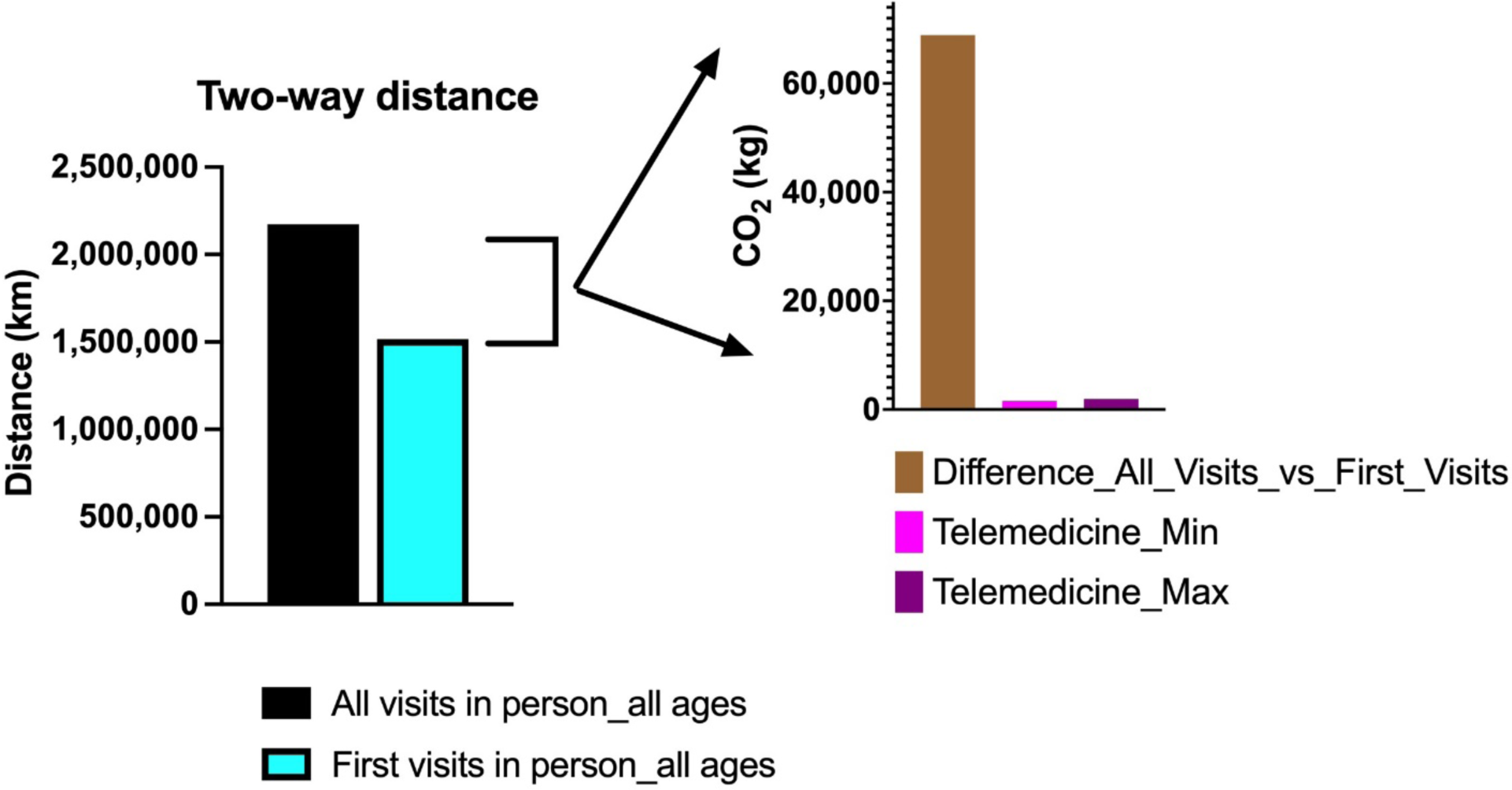
The distance and CO2 difference if the first visits are done in person with subsequent televisits.

### 3.2. Carbon footprint

In this study, calculations of carbon footprint were made to show the effects of hospital visits on the environment. Access to the hospital can be possible in various ways. However, to simplify the calculations, it is assumed that one can travel either by bus or car.

The carbon footprint of traveling by bus is 105 grams per kilometer and the carbon footprint of traveling by car is 192 grams per kilometer. Since it is necessary to make round-trip calculations in carbon footprints, the figure found was multiplied by two. For example, if the distance from the address of the patient to the hospital is 10 kilometers:

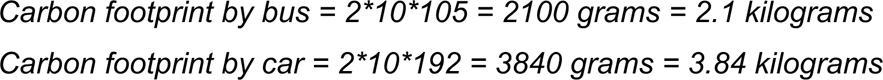

From the above calculations, it is seen that the cost of traveling by car is 82.86% higher than traveling by bus. On the other hand, if the patient in the example does not go to the hospital and is examined remotely, the footprint is minimal.

The digital footprints were calculated for a ten-minute doctor-patient video conferencing, and it is assumed that on the average one large image is exchanged for diagnosis. The digital footprint is independent of the distance between the hospital and the address of the patient. The calculations are as follows:

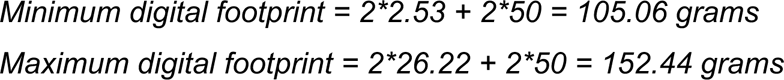

In min (max) digital footprint calculation, a ten-minute video conference cost is taken as 2.53 (26.22) grams and is multiplied by two to account for both the doctor and the patient. The added term is constant, and it is the cost of sending and receiving one large image file. It is assumed that the footprints of sending an image file and receiving it are approximately equal. The footprint values in the calculations except for the image are taken from Obringer et al. [29]. On the other hand, the footprint of an image is taken from Frost [30]. The dataset in this study contains 45,602 visits to the hospital and the total distance of the roundtrips by the patients turns out to be 2,173,262 kilometers. If all the roundtrips are made by bus, then the total amount of carbon footprint is 228,192.61 kilograms for all in-person visits, whereas it could decrease to 159,285 kilograms if the first visits were done in person with subsequent visits via telemedicine (Fig. 8).

**Figure 8:**
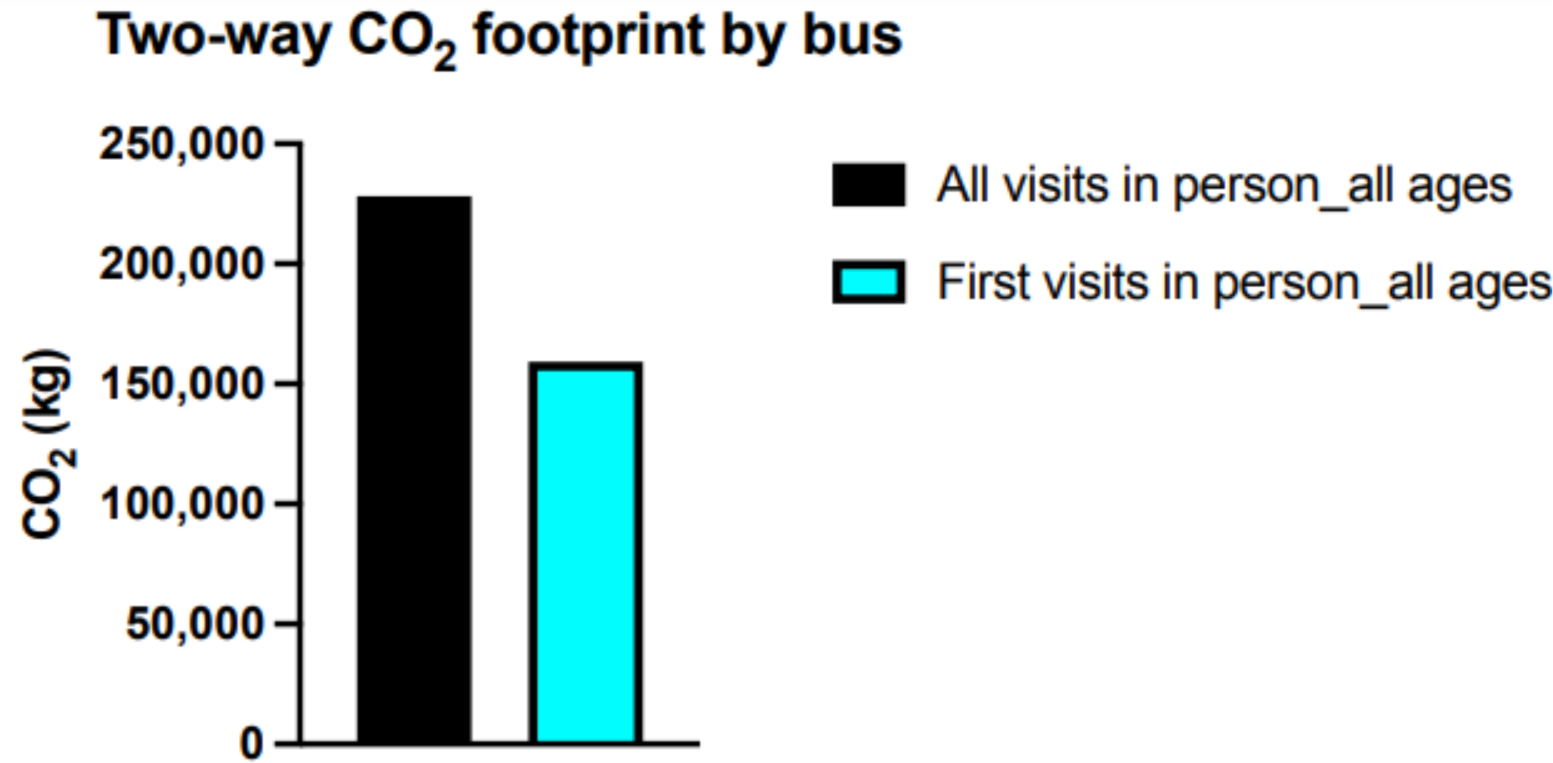
CO2 footprint difference for public transportation if the first visits are done in person with subsequent televisits.

Similarly, if all visits are made by car, then the total amount is 417,266.49 kilograms, which would decrease to 291,264 kilograms if the first visit were done in person with subsequent visits via telemedicine (Fig. 9). The real amount is expected to be in the range of 159 to 417 tons.

**Figure 9:**
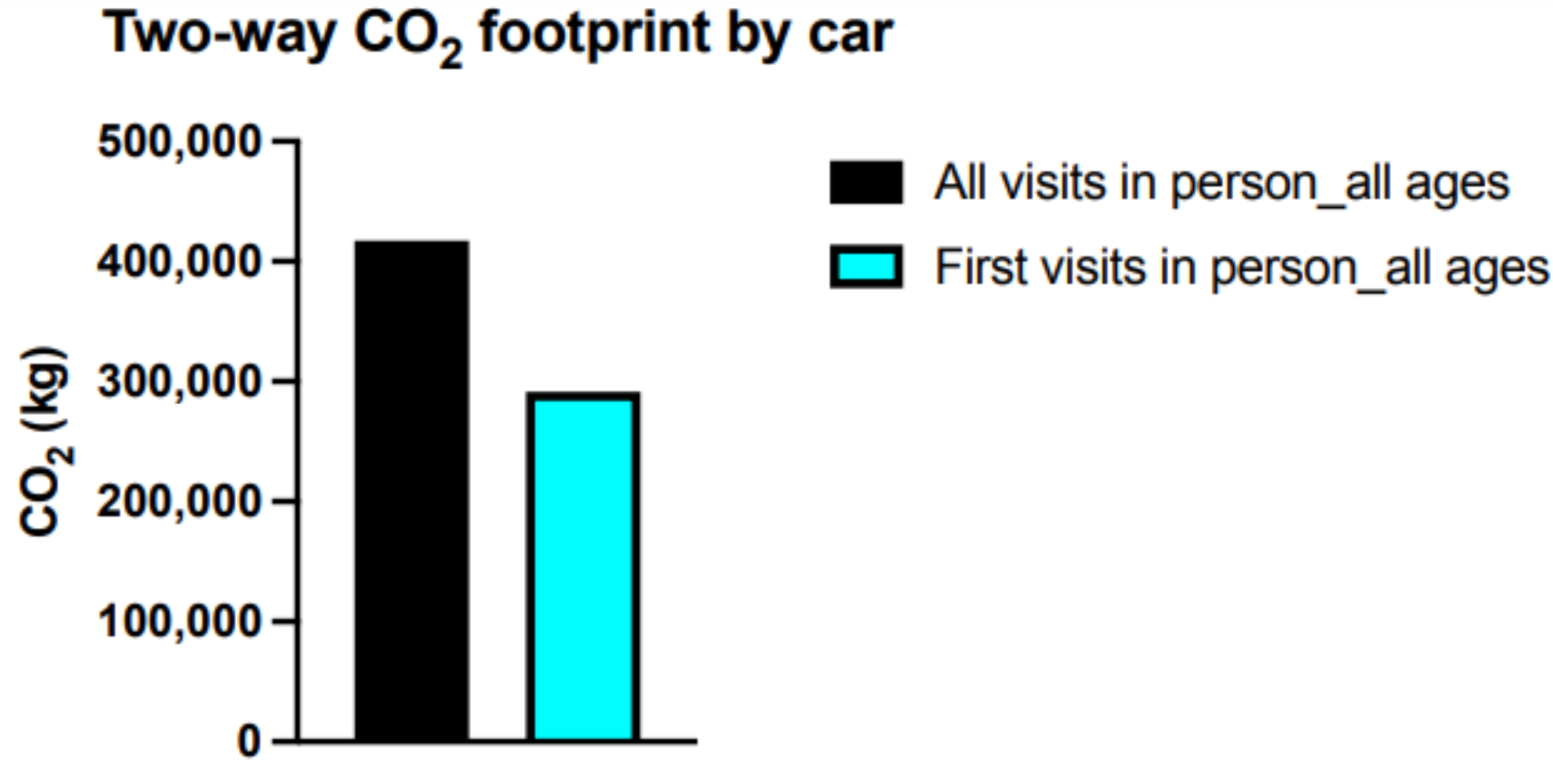
CO2 footprint difference for personal vehicle transportation if the first visits are done in person with subsequent televisits.

Our assumption that all but first in-person visits could be replaced with telemedicine without compromising quality of patient care relies on the fact that, for the diagnoses that we selected, 98.9% of the visits subsequent to the first were follow-up visits that did not require any in-person intervention. This is also in line with the opinions of the cardio-vascular specialists working at these facilities we consulted while conducting our study.

In this scenario, the total distance that must be traveled is 1,517,004.92 kilometers, which means a 30.20% decrease in the total amount of distance to be traveled. In this scenario, the number of visits is 29,985, in other words 34.25% less than the ‘all visits in person’ scenario. The effect on carbon footprint is also dramatic. If the first visits are done by bus, the difference of the carbon footprint is 68,907.10 kilograms. On the other hand, if the first visits are done by car, the difference of the carbon footprint is 126,001.55 kilograms. In both cases, a 30.20% reduction in the carbon footprint is achieved (Fig. 8 and 9).

### 3.3. Wage loss

Wages were derived from the ILO site for the study period for Turkey and the average wage loss is calculated by multiplying the number of visits to the total time spent for traveling and visits. For the physician visits, a 10-min time is allotted. The 2021 average hourly wage for Turkey was 28.13 Turkish Lira (TL), where one USD was worth 8.86 TL in 2021, making the average hourly wage $3.26 [31]. Table 1 shows the travel times with an additional 10-min for in-person visit for working-age males and females in our cohort. The wage lost is presented based on the time spent traveling and in-person visit (“Wage loss per hour”) as well as a whole day loss of productivity based on the number of visits. Additionally, these travel times are calculated based on the use of a personal vehicle, where the use of public transportation would increase the travel times.

**Table 1:** Wage loss caused by in-person hospital visits.

|  | **Males < 60 years** | **Females < 58 years** |
| --- | --- | --- |
| **Total visits** | 13,995 | 19,656 |
| **Travel distance (km)** | 622,299 | 755,425.8 |
| **Travel times (hr.)** | 15,812.6 | 18,782.7 |
| **Each visit with added 10 min for in-person travel times (hr.)** | 22,810.1 | 28,610.7 |
| **Wage loss for an 8-hr. day per visit (\$)** | 364,989.6 | 512,628.48 |
| **Wage loss per hr. (\$)** | 74,360.9 | 93,270.8 |

## 4. Discussion

The global carbon footprint of health care was estimated to be 2 gigatons of CO_2_-equivalents (CO_2_e) in 2014, approximating 4.4% of global emissions, whereas global transportation contributed 7% of this total. Thus, in a world where climate change is a major risk for human health, the health care continues to significantly contribute to this hazard [32], [33]. Along the environmental pressures of delivering health care, to deliver it in a sustainable fashion is a growing concern for every country because of both increasing costs and the increasing ratio of health expenditure to gross domestic product (GDP) [34]. For example, in Turkey, in the two decades until 2020, health care expenditure increased from $432/capita in 2000 to $1,305/capita in 2020 [35], nearly tripling in 20 years and the ratio of health expenditure to GDP increased from 4.46% to 5% [36], [37]. This is not an isolated phenomenon and between 2000 and 2020 as the worldwide health expenditure to GDP increased from 8.62% to 10.89% [38].

In addition to increasing health care cost, each visit to a health care facility results in increased carbon emissions due to travel and loss of productivity. The travel-related carbon emission within the context of global warming is an important variable that is integral to a more sustainable health care system. A recent cross-sectional study by Patel evaluated 49,329 telemedicine visits by 23,228 patients to the Moffitt Cancer Cancer (MCC) from April 1, 2020 to June 30, 2021. The authors divided the telemedicine cohort into two groups based on travel times (<60 and >60 min) and calculated the associated CO_2_ emission savings. In their cohort, patients living within a driving distance of 60 minutes from the cancer center saved on average 19.8 kg CO_2_ emissions per-visit due to telemedicine. And for patients whose driving were for more than 60 minutes, 98.6 kg CO_2_ emissions was saved per visit. The majority of the patients coming to MCC were within 60 minutes of one-way driving time [39]. While their work and results align with our findings, the scales are different as the population density is approximately **18.5 times higher** in Istanbul, Turkey (2,523 people/km^2^ or 6,530 people/mi^2^) compared to Florida, USA (136.4 people/km^2^ or 353.4 people/mi^2^), which could explain the longer travel distance and time per visit in Florida [40], [41].

The telemedicine technology can be grouped according to type of *interaction* (clinician-to-patient or clinician-to-clinician) or *timing* (asynchronous or synchronous). Asynchronous involves sending pre-recorded information between individuals, whereas synchronous is real-time data transmission. The data may be transmitted via a variety of media, such as audio, video or text.

Telemedicine is a recent development within healthcare. The reported advantages include lower healthcare costs, high patient satisfaction, improved access, decreased wait times and fewer missed appointments. The purported disadvantages are depersonalization of the clinician–patient relation and concerns around quality of care. Despite the extensive effectiveness research, cost and perceptions of telemedicine, there have been few contributions assessing the environmental impact, such as travel times [24], [42]–[44].

Telemedicine could help to contain the increasing health care costs in a sustainable way if used in appropriate settings and accepted by the society. We therefore attempted to define the impact of the current health care delivery model of in-person visits to a busy health care system in Istanbul, Turkey, with the goal of developing a framework of a cost-conscious and sustainable model [45]. Based on our data, the average person did visit the clinics from a distance of 47.6 ± 96.3 km and spent 67.2 ± 108.9 min travel time and associated CO_2_ emission 4,998 (bus)/ 9,139 gr (car).

Similar to the CO_2_ emission, the loss of productivity also impacts the sustainability. The average wage loss was $4.20 ([two-way travel time + 10 min physician time (hr)]*$3.26). Our data suggests that using telemedicine would lead to significant cost savings as well as a positive impact on the environment. In addition, the existence of a single payor system in Turkey brings advantages to test the telemedicine model on a larger scale.

## 5. Limitations

While it is exciting and promising to use telemedicine in delivering quality care to patients, there are several limitations for our work.

First of all, we used a data from a very specialized healthcare system serving patients with cardiovascular diseases and this may not be applicable to other healthcare systems.

Secondly, the cost-effective interventions could be unaffordable [46]. The cost-effective benefit may not be achieved if the implementation (internet infrastructure, servers, end-user devices and applications) is prohibitively expensive [47]–[49]. In addition, the few programs that start successfully may not always be sustainable in the long run, scalable, or transferable to other settings [50].

As a third limitation, the acceptance by patients and providers is another unknown in the application of such technologies. During the COVID-19 pandemic, the use of telecare has been well accepted by both the clinicians and the patients in Turkey [26], [51]. For our data calculations, we made assumptions of secondary visits to be performed over video platforms and supported this assumption by the expert cardio-vascular specialists, however the acceptance among patients is not clear.

Finally, as for the travel time calculations, we relied on the Google Maps algorithms and used the after-hours travel, however the traffic patterns may change and that negatively impact our time calculations and increase the time spent traveling. We defined these travel times as the “floor” times and these results should only be treated as the minimum possible travel time. In addition, the wait times during in-person visits are omitted from our calculations for the wage loss as we did not have the data for the wait times, and this could be a future direction of our work.

## 6. Conclusion

In this paper, we examined the potential impact of delivering tele-healthcare to patients by using a dataset that spans more than seven years from a tertiary cardio-vascular clinic in Istanbul, Turkey. We estimated the carbon footprint of traveling for in-person visits and the incurred wage loss due to time spent for these visits. The results presented in this paper indicate that application of telemedicine in Turkey could provide a feasible solution to decrease the burden on patients, environment, increase access, and prevent wage loss caused by unnecessary hospital visits. This way, telemedicine could become a valuable tool for sustainable cities of the future.

## Data Availability

The address data will not be available.

## Footnotes

2 The figure is derived from (https://www.healthdata.org/turkey), accessed 1 June 2023.

